# Phenotype correlations with pathogenic DNA variants in the *MUTYH* gene

**DOI:** 10.1101/2024.05.15.24307143

**Authors:** Monica Thet, John Paul Plazzer, Gabriel Capella, Andrew Latchford, Emily AW Nadeau, Marc S Greenblatt, Finlay Macrae

## Abstract

*MUTYH*-associated polyposis (MAP) is an autosomal recessive disorder where the inheritance of constitutional biallelic pathogenic *MUTYH* variants predisposes a person to the development of adenomas and colorectal cancer (CRC). It is also associated with extracolonic and extraintestinal manifestations that may overlap with the phenotype of familial adenomatous polyposis (FAP). Currently, there are discrepancies in the literature regarding whether certain phenotypes are truly associated with MAP. This narrative review aims to explore the phenotypic spectrum of MAP to better characterise the MAP phenotype. A literature search was conducted to identify articles reporting on MAP-specific phenotypes. Clinical data from 2109 MAP patients identified from the literature showed that 1123 patients (53.2%) had CRC. Some patients with CRC had no associated adenomas, suggesting that adenomas are not an obligatory component of MAP. Carriers of the two missense founder variants, and possibly truncating variants, had an increased cancer risk when compared to those who carry other pathogenic variants. It has been suggested that somatic G:C>T:A transversions are a mutational signature of MAP, and could be used as a biomarker in screening and identifying patients with atypical MAP, or in associating certain phenotypes with MAP. The extracolonic and extraintestinal manifestations that have been associated with MAP include duodenal adenomas, duodenal cancer, fundic gland polyps, gastric cancer, ovarian cancer, bladder cancer and skin cancer. The association of breast cancer and endometrial cancer with MAP remains disputed. Desmoids and Congenital Hypertrophy of the Retinal Pigment Epithelium (CHRPEs) are rarely reported in MAP, but have long been seen in FAP patients, and thus could act as a distinguishing feature between the two. This collection of MAP phenotypes will assist in the assessment of pathogenic *MUTYH* variants using the American College of Medical Genetics and the Association for Molecular Pathology (ACMG/AMP) Variant Interpretation Guidelines, and ultimately improve patient care.

## 1. Introduction

Gastrointestinal polyposis syndromes are characterized by tens to thousands of polyps, of which adenomatous polyposis is the most common type. Adenomatous polyps are precancerous lesions which, if undetected, can develop into colorectal cancer (CRCs). To date, numerous types of inherited adenomatous polyps have been discovered. While each inherited form harbours adenomatous polyps, the quantity of polyps and the extracolonic manifestations can vary depending on the type.

*MUTYH*-associated polyposis (MAP) is one of these adenomatous polyposis syndromes. First described in 2002 by Al Tassan et al., it is an autosomal recessive disorder where one inherits constitutional biallelic pathogenic variants in the *MUTYH* gene (1). These pathogenic variants predispose individuals to the development of adenomatous polyps and CRCs (1-5). *MUTYH* is one of the base excision repair (BER) genes located on chromosome 1 (1p34.3-p.32.1) and is involved in the repair of oxidative damage. The two founder variants, p.(Tyr179Cys) and p.(Gly396Asp), are by far the most common pathogenic *MUTYH* variants in Caucasian populations (1, 4-7). Because of alternative start codons and alternative splicing, the amino acid positions of these codons have not been agreed upon in the literature. The most commonly cited transcript corresponds to p.Tyr179Cys and p.Gly396Asp, with allele frequencies in European populations of 0.15% and 0.3%, respectively, according to the ClinVar database at the National Centre for Biotechnology Information (NCBI).

Constitutional pathogenic *MUTYH* variants account for 30-40% of cases with adenomatous polyposis where a constitutional variant in the adenomatous polyposis coli (*APC*) gene is not detected (4-7). There are also reported cases where polyps are absent (8-10), and therefore the term MAP may not represent the entire population of patients with constitutional biallelic pathogenic *MUTYH* variants. “*MUTYH*-associated tumour syndrome,” an alternative to MAP, has been proposed to better represent the broad phenotypic range associated with pathogenic *MUTYH* variants (11).

MAP is also associated with several extracolonic and extraintestinal manifestations, and these may overlap with, but not duplicate, the phenotype of familial adenomatous polyposis (FAP) (12). A comprehensive characterisation of the MAP phenotype will improve the diagnosis and treatment of MAP. Moreover, it will also contribute to the development of *MUTYH*-specific American College of Medical Genetics and the Association for Molecular Pathology (ACMG/AMP) Variant Interpretation Guidelines, which will standardize the characterization of *MUTYH* variants. This narrative review explores the phenotypic spectrum of MAP described in the literature, including colonic, extracolonic and extraintestinal manifestations, and aims to better describe the syndrome-specific phenotype of *MUTYH*-associated tumour syndrome.

## 2. Methods

To find articles describing the phenotypes of biallelic pathogenic *MUTYH* variants, the following key search terms were used: MUTYH, MUTYH-associated polyposis, base excision repair, adenomatous polyp, colorectal cancer, colorectal adenoma, gastrointestinal cancer, extracolonic cancer/tumour, extraintestinal cancer/tumour, phenotypic variability, and genotype-phenotype correlation (See Supplemental Section S1 for search strategy and results). All relevant references within these articles were also included. The following exclusion criteria were applied: review articles prior to 2018 (except those that are highly cited in the literature), non-English articles, articles that are out-of-date or without data of interest, irrelevant populations (monoallelic or healthy controls) and articles looking at interactions between *MUTYH* and other genes (Figure 1). The cut-off year of 2018 was chosen because comprehensive reviews for *MUTYH* published around and after 2018 also collected relevant information from studies conducted prior to 2018.

**Figure 1.**
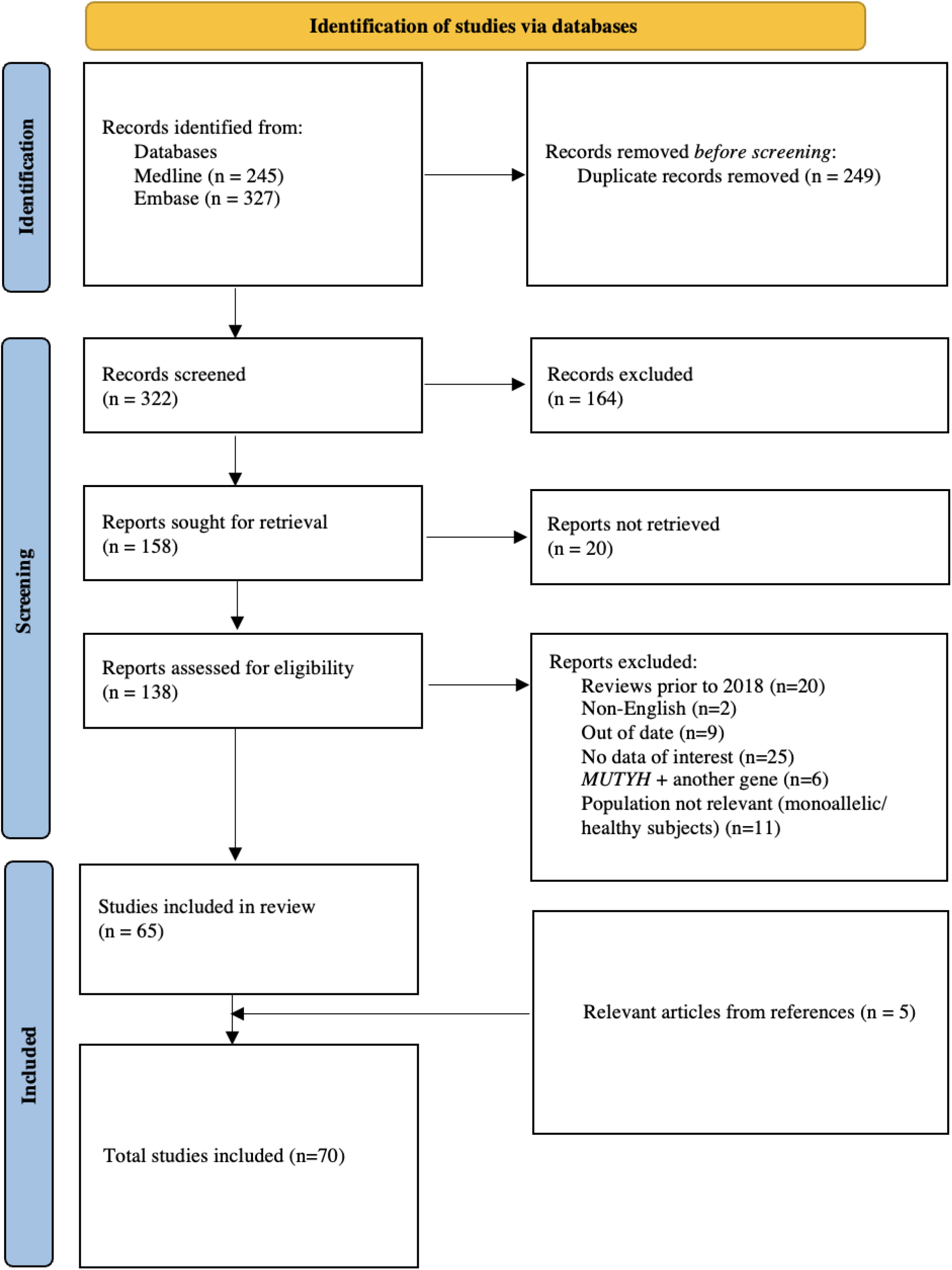
PRISMA flow chart illustrating the systematic approach for study inclusions through the stages of this narrative review.

Pathogenic *MUTYH* variants from the literature, ClinVar and the Leiden Open Variation Database (LOVD) were mapped to the gene in Figure 2. A five-tier system is currently used to classify variants into one of the following categories: Pathogenic, Likely Pathogenic, Variant of Uncertain Significance (VUS), Likely Benign, and Benign. If a variant had been classified under more than one category, the higher classification was used. For instance, if a variant was classified as both Pathogenic and Likely Pathogenic, it was considered as Pathogenic and thus included in Figure 2. The reference transcript NM_001128425.2 was used to describe pathogenic variants.

**Figure 2.**
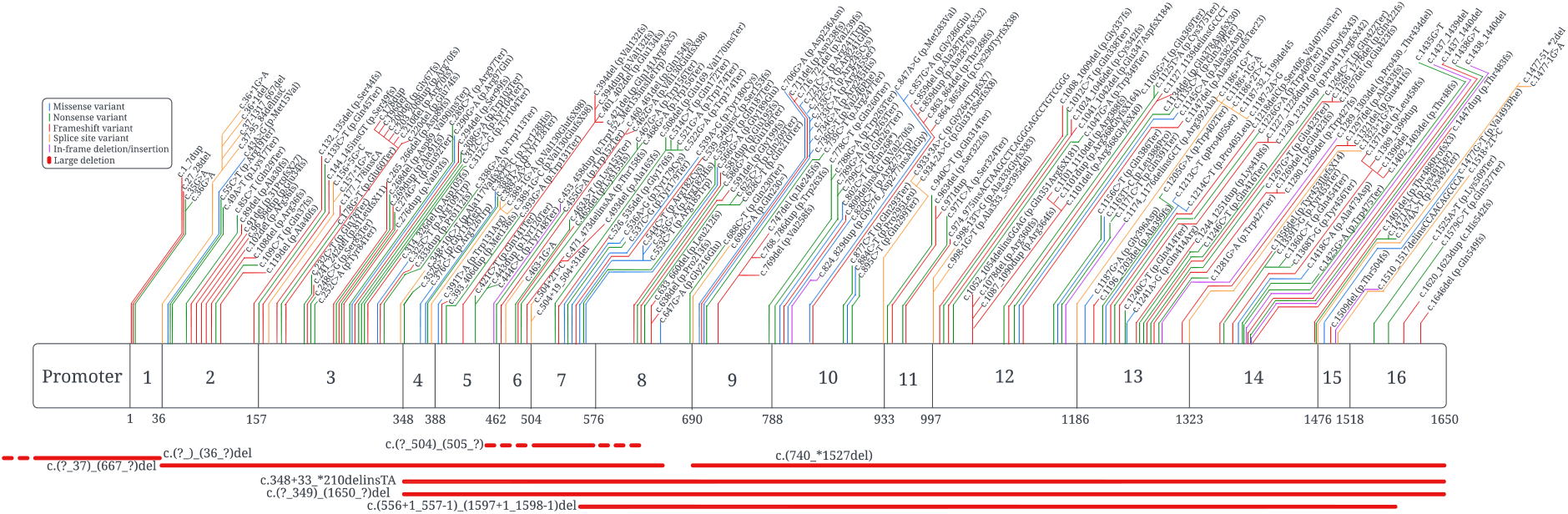
Location and type of Pathogenic variants in *MUTYH* identified in the literature, ClinVar and LOVD.

## 3. Results and Discussion

A total of 234 pathogenic variants were identified in the literature, ClinVar and LOVD. Of these, 77 were reported in articles describing the phenotypic characteristics of the affected individuals. The remaining 156 variants were not identified in the literature but were reported in ClinVar, LOVD, or in both ClinVar and LOVD. Frameshift (n = 87) and nonsense (n = 71) variants were the most common types of pathogenic *MUTYH* variants (Table 1). Pathogenic missense variants can occur throughout the entire length of the protein and are not confined to one region or domain.

**Table 1.** Frequency of each variant type reported in Figure 2.

| Variant Type | Number Reported in Literature, LOVD and ClinVar | Number Reported in Literature with Phenotype |
| --- | --- | --- |
| Missense | 33 | 29 |
| Nonsense | 71 | 16 |
| Frameshift | 87 | 14 |
| Splice site | 27 | 10 |
| In-frame insertion/deletion | 7 | 6 |
| Large deletion | 9 | 2 |
| Total | 234 | 77 |

Clinical features of MAP patients reported in the literature are summarized in Table S2. Population inclusion criteria varied across the studies. However, patients with adenomas or CRCs who had a pathogenic variant in *APC* and/or mismatch repair (MMR) genes were consistently excluded. Variant analysis was conducted in one of two ways. The first method involved sequencing the entire coding region, with or without exon-intron junctions, to identify variants. The second method involved first sequencing the two pathogenic founder variants, p.(Tyr179Cys) and p.(Gly396Asp). Patients who were heterozygous for one of these two variants then had the entire *MUTYH* gene sequenced to search for a second pathogenic allele. It should be noted that this latter method has a possibility of missing MAP patients with non-founder pathogenic variants.

The identification of extracolonic and/or extraintestinal manifestations was dependent on the degree of intensity that they were looked for and as such, not all studies reviewed here included findings about these manifestations. This applies particularly to the index cases in families who are generally symptomatic when compared to other affected family members, and hence were subject to more extensive investigation. Overall, the percentage of MAP patients with upper gastrointestinal (GI) findings was calculated based only on the population of patients in the study who underwent gastroscopy. Of the 2109 MAP patients recorded in Table S1, 1123 patients had CRC (53.2%). 936 patients underwent upper GI gastroscopy and 160 of these patients were reported as having duodenal adenomas (17.1%). Three patients had duodenal cancer (0.3%), all of whom had concurrent duodenal adenomas at the time of diagnosis. Summary statistics were not reported for other phenotypes due to incomplete investigation across the studies.

### 3.1 Variant pathogenicity

In Western populations, up to 70% of MAP patients carry the two pathogenic founder variants, p.(Tyr179Cys) and p.(Gly396Asp), in either homozygosity or compound heterozygosity. Moreover, as many as 93% of MAP patients carry at least one of these two variants (6). Other pathogenic *MUTYH* variants that may be specific to certain ethnic groups include p.(Tyr104Ter), p.(Glu466Ter), p.(Glu480del) and c.1227_1228dup p.(Glu410GLyfsX43) in Pakistani, Indian, Italian and Portuguese populations, respectively (4, 13-15). The variant c.1227_1228dup has also been found in high frequency in North African patients (16-18).

Due to the widely available data for these two founder variants in Western populations, multiple studies have evaluated their pathogenicity. Studies have found that p.(Tyr179Cys) homozygous carriers had earlier disease onset, more severe phenotype and increased cancer risk when compared to homozygous carriers of p.(Gly396Asp) and compound heterozygous carriers of these two founder variants (9, 10, 19-21). The risk of colorectal cancer was doubled in patients carrying these two founder variants, either in homozygosity or compound heterozygosity, when compared to biallelic carriers of other pathogenic variants (10).

Evidence suggests that the molecular consequences of variants also have varying pathogenicity. Patients with truncating pathogenic variants and in-frame deletions that result in exon skipping have been reported to have a more severe phenotype than patients with missense pathogenic variants (19, 22, 23). In a study by Nielsen et al. (2009) that contained 257 MAP patients, more patients with biallelic truncating pathogenic variants (5 of 11, 45%) appeared to have polyp counts of >100 in comparison to those with zero (28 of 109, 26%) or one (4 of 28, 15%) truncating pathogenic variants, even though this finding was not statistically significant (19). Interestingly, the same study also found that none of the patients with two truncating pathogenic variants had fewer than 10 polyps.

Based on this data, it can be inferred that different pathogenic variants result in varying degrees of pathogenicity. Patients with the same pathogenic variant may have varying numbers of polyps, may or may not develop CRC, and may or may not develop other extracolonic phenotypes (6, 7). This suggests that phenotypic heterogeneity is not governed only by specific pathogenic *MUTYH* variants. Instead, the penetrance of these variants is likely influenced by epigenetic, other genetic and environmental factors (24).

### 3.2 Age of diagnosis

The reported age of diagnosis of patients with MAP varied across the studies included in this review, with the median age at diagnosis ranging from 45 to 58 (Table S1). The youngest case reported was a 13 year-old Caucasian male, compound heterozygous for p.(Tyr179Cys) and p.(Gly396Asp) (2). He had more than 100 polyps at the time of diagnosis. No CRC was diagnosed at the time, but he developed gastric cancer at the age of 17. Based on the recruitment criteria of this study (2), he had no pathogenic *APC* variant identified and there was no vertical transmission of polyposis in his family. However, his age at diagnosis is unusual and lies well outside the median age range at diagnosis for biallelic *MUTYH* patients, suggesting that there may be other factors contributing to the case.

### 3.3 Somatic G:C>T:A transversions

In the presence of reactive oxygen species, guanine is converted to 8-oxoguanine (8-oxoG), a compound that mis-pairs with adenine. The *MUTYH* gene encodes a 535 amino acid DNA glycosylase that identifies and removes adenine bases mis-paired with 8-oxyguanine (1). However, if these A:8-oxoG mis-pairs are left incorporated in DNA, they lead to somatic G:C>T:A transversions upon replication. These transversions are typically observed in the *APC* gene in adenomas of people with constitutional, biallelic, pathogenic *MUTYH* variants (6, 13, 25-27), as well as in other genes such as *KRAS* (13, 25-28).

At least ten different nucleotide substitutions, predominantly in codons 12, 13 and 61, are prevalent in multiple tumours and colorectal cancer (29). However, only two of these G:C>T:A transversions are at the first (base 34, results in Gly12Cys) and second (base 35, results in Gly12Val) nucleotides of codon 12. These transversions are oncogenic and can frequently occur in both FAP-associated colorectal tumours and sporadic tumours (26, 30). Interestingly, *KRAS* mutations in MAP tumours are exclusively G:C>T:A transversions at the first G of codon 12 (25, 26, 28, 30, 31). The finding of this specific somatic Gly12Cys variant in the *KRAS* gene in all observed patients with constitutional biallelic pathogenic *MUTYH* variants compared to patients with sporadic (13.6%) or FAP-associated adenomas or CRCs (18.5%) is statistically significant (p≤0.001 and p≤0.002, respectively) (13, 26). Another recent study that compared the prevalence of somatic mutations in colorectal cancers of patients with constitutional biallelic pathogenic *MUTYH* variants (n = 19) to patients without constitutional or somatic pathogenic *MUTYH* variants (n = 5364) found that patients with *MUTYH* variants had a significantly higher proportion of somatic *KRAS* G>T transversions at codon 12 (Gly12Cys) (84% vs. 2.4%; p = 2.0×10^-23^) (32). Therefore, this variant could act as a potential biomarker for screening and identifying patients with atypical MAP, *i*.*e*., without polyps or with less than 10 polyps (31). Moreover, screening for this variant in extracolonic tumours of MAP patients could help differentiate between tumours that occur sporadically and those that are caused by pathogenic *MUTYH* variants, which allows for a more accurate representation of the MAP phenotypic spectrum. This biomarker could also be used to identify an additional cohort of *MUTYH* carriers whose families would benefit from genetic counselling.

### 3.4 Colorectal cancer and colonic polyps

MAP patients have an increased risk of developing CRC compared to the general population, with approximately two thirds of MAP patients developing CRC (3, 6, 33, 34). MAP patients have variable age of diagnosis, polyp count, polyp type, and may have CRC at different locations. CRC is more commonly confined to the right side of the colon, with at least 60% of MAP patients having CRC proximal to the splenic flexure (3, 19, 28, 35-37). The mean age of CRC diagnosis in MAP patients is 42.3-50 years, and 95% of patients are diagnosed with CRC after the age of 35 (3, 38-40). The age at diagnosis of CRC in MAP patients is younger than patients with sporadic CRC (68yr) (28, 38, 41). The youngest reported CRC patient thus far is a 21 year-old female with 36 polyps and compound heterozygous for p.(Tyr179Cys) and c.1147del p.(Ala385ProfsTer23) (3).

Most colonic polyps in MAP patients are adenomas, and polyp counts are variable. It has been found that MAP patients have a higher polyp count and earlier presentation (median age = 47yr) compared to non-MAP patients (median age = 54yr) (40). While some studies have suggested that it is rare to only have a few adenomas in patients with biallelic pathogenic *MUTYH* variants (2-4, 6, 19), polyps may not be an obligatory phenotype of MAP (10), as the absence of polyps has been noted in multiple studies (8-10, 34). However, studies that mentioned the absence of polyps were conducted prior to 2010. Since that time, there have been significant advances in endoscopy (high-definition imaging, narrow band imaging, artificial intelligence, etc.) that have facilitated polyp detection. Therefore, if phenotype was ascertained through colonoscopy, it is possible that polyps may have been overlooked due to older endoscopic technologies and these patients may have had a more typical MAP phenotype. Patient cohorts that have polyp counts between 10-99 have been reported to have more carriers of pathogenic *MUTYH* variants when compared to patient cohorts that have a polyp count between 100-1000 (2, 4, 6, 25). However, other studies have found that pathogenic, biallelic *MUTYH* variants occur at equal frequencies in both patient groups (3, 42). These differences could be due to different methods of ascertainment, including symptomatic index cases and asymptomatic relatives. Although not included in our review, phenotype is likely to be more variable in cohorts identified by panel testing of phenotypes not associated with polyposis or colorectal cancer. In addition to high polyp count not being a necessary component of MAP (10), polyp count may also not be a reliable predictor of CRC development. Several studies have suggested that high polyp burden is not associated with the occurrence of CRC (41, 43). In fact, colorectal cancer has developed in patients with fewer than 10 adenomas (41). These contrasting reports may indicate that polyp count and the development of CRC are not reliable indicators for genetic testing aimed at identifying *MUTYH* variants.

The most frequent polyp type described in MAP is tubular adenomas. Tubulovillous adenomas and, rarely, hyperplastic polyps (HP) and sessile serrated lesions (SSL), have also been reported (20, 44-46). Boparai et al. (2008) found that 47% (8/17) of patients had HP and SSL in addition to adenomas. While adenomas in this study harboured both *APC* and *KRAS* mutations with somatic G:C>T:A transversions, HP and SSL only had *KRAS* mutations (44). G:C>T:A transversions in HP and SSL were more often observed in MAP patients (51 of 73 patients) than the control group with sporadic adenomas (7 of 41 patients; p<0.0001). Data suggests that these polyp types may also be a part of the MAP phenotype. Alternatively, there could be a subset of serrated polyposis syndrome where *MUTYH* is a contributing factor but not the root cause (47). For example, an unknown interaction between *MUTYH* and another gene could be the cause.

### 3.5 Extracolonic GI manifestations

Although data on the presence of somatic G:C>T:A transversions in the *KRAS* gene in duodenal lesions in MAP patients is limited (48), by analogy with the colon, the presence of somatic G:C>T:A transversions in duodenal lesions would strongly support the likelihood that duodenal lesions are caused by a deficiency in *MUTYH* activity (48). The prevalence of duodenal polyps ranged from 17% (26/150) of the cases in Vogt et al. (2009), 18% (6/33) of the cases in Aretz et al. (2006) and 34% (31/92) of cases in Walton et al. (2016) (5, 12, 49). This variability in prevalence may be because not all MAP patients in these studies underwent endoscopic examination of the upper GI tract. The age at the time of the endoscopic evaluation also varied. Therefore, these estimates concerning the prevalence of duodenal polyps in MAP remain preliminary. It should also be noted that the histopathological characteristics of duodenal lesions were not always described. However, outside of the hamartomatous polyposis syndromes, duodenal polyps other than adenomas are exceedingly rare. In the three studies described above, all 31 patients with duodenal polyps in Walton et al. (2016) and 16 of 26 patients in Vogt et al. (2009) had histologically confirmed adenomas, and 1 patient had hyperplastic polyps. The polyp type was unknown in the remaining 9 patients. Polyp type was not described in Aretz et al. (2006).

When assessed using Spigelman staging, the prevalence and severity of duodenal adenomas is lower in MAP than in FAP (12, 27, 49, 50). The lifetime risk of developing duodenal cancer in MAP is not well established, but it has been reported to be 4%, which is higher than the general population risk (12). While Spigelman stage IV is a strong predictor of duodenal cancer development in FAP, this does not seem to be the case for MAP (48-50). In contrast to patients with FAP, MAP patients have far fewer duodenal adenomas, and many patients have a solitary adenoma (49, 50). Furthermore, in MAP, progression through the Spigelman classification occurs predominantly based on progression of histological features and not by progression of adenoma count (49, 50). It has also been found that, despite having lower Spigelman stage, duodenal adenomas associated with MAP have a higher burden of somatic variants in oncogenic driver genes such as *APC* and *KRAS* when compared to FAP-associated adenomas (27). In a study that performed whole exome sequencing on 10 duodenal adenomas in 5 MAP patients and 10 duodenal adenomas in 4 FAP patients, the rate of *APC* somatic G>T transversions in MAP adenomas (481/716; 67%) was higher than in FAP adenomas (28/225; 12%) (p<2.2×10^-16^). MAP duodenal adenomas also had significantly more *KRAS* mutations than FAP adenomas (8/22 vs. 4/38 respectively; p<0.023) (32). This higher mutation burden could increase duodenal cancer risk in patients with MAP despite the manifestation of duodenal adenomas (number, size, etc.) being less severe. These observations suggest that it may not be appropriate to use Spigelman staging in MAP-associated duodenal adenomas to predict future disease progression. As such, a low duodenal adenoma count may be falsely reassuring to MAP patients (27).

The risk of developing duodenal adenomas in MAP may be associated with specific pathogenic variants. In a multi-centre study involving endoscopic surveillance of the duodenum of 394 MAP patients, patients homozygous for the variant p.(Tyr179Cys) were more likely to have adenomas at initial endoscopy when compared to p.(Gly396Asp) homozygotes or p.(Tyr179Cys)/p.(Gly396Asp) compound heterozygotes (50). Moreover, adenomas in p.(Tyr179Cys) homozygous carriers were more likely to harbour high-grade dysplasia and/or contain villous components (8 of 67, 12%) when compared to the latter two (0 of 62 p.(Gly396Asp) homozygotes; 2 of 55 p.(Tyr179Cys)/p.(Gly396Asp) compound heterozygotes). This finding further supports the above observation that p.(Tyr179Cys) homozygous carriers have a more severe phenotype than others (19).

There is no robust association between gastric cancer and MAP. However, gastric cancer in MAP patients has been reported in some studies and has also been reported in *MUTYH* heterozygotes (2, 12, 36, 51, 52). It may well be that there were additional factors that influenced the early development of gastric cancer in these cases (2). Fundic gland polyps are also described in MAP literature, although whether they are observed more commonly than the general population is not clear (12, 38, 53). The occurrence of pyloric gland adenomas in MAP has only been reported once, although it has been reported in about 6% of FAP patients (54).

### 3.6 Extraintestinal manifestations

Many extraintestinal manifestations have been found in MAP patients. However, the extent to which pathogenic *MUTYH* variants are directly responsible for or are associated with these manifestations is not clear. Although controversial, there is some evidence for the associations outlined below, and thus they are potentially important components of the MAP phenotypic spectrum. This dispute is due to the lack of statistically significant extraintestinal manifestations in MAP patients when compared to the general population.

In a retrospective study that involved 276 patients, the incidence of three extraintestinal malignancies was almost doubled in MAP patients when compared to the general population (12). The incidence for ovarian (standardised incidence ratio [SIR] = 5.7; 95% confidence interval [CI] = 1.2–16.7), bladder (SIR = 7.2; 95% CI = 2.0–18.4) and skin cancers (SIR = 2.8; 95% CI = 1.5-4.8) were increased significantly. It is important to note that the population in this study not only included the index patients and their affected relatives with biallelic pathogenic *MUTYH* variants, but also their deceased relatives who had confirmed colorectal adenomatous polyps and/or identified pathogenic, biallelic *MUTYH* variants. This selection criteria suggests that the deceased relatives with polyps, who were not genetically tested, might have been included in the study. Because their phenotype was not confirmed via genotype, their inclusion could have led to ascertainment bias. Nevertheless, some of the findings of this study were supported by Win et al. (2016), where the risk of bladder cancer and ovarian cancer were increased by 19-fold and 17-fold, respectively (55).

The association between breast cancer and *MUTYH* is also contested (3, 12, 55, 56). In one study, 4 of 22 female Dutch MAP patients developed breast cancer, two under age 50. No study has replicated this observation and thus confirmed a significantly increased risk of breast cancer in biallelic *MUTYH* carriers (3). Moreover, in another study looking at the prevalence of *MUTYH* mutations in 691 breast cancer patients and 812 healthy controls, it was found that none of the participants carried the two founder mutations (56). However, this study only screened for the two founder mutations, and did not sequence the entire gene. Thus, this study could have failed to identify other pathogenic *MUTYH* variants. As such, more studies are required to fully explore the association between the risk of breast cancer and the presence of biallelic *MUTYH* variants.

Endometrial cancers have also been reported in MAP patients, but like breast cancer, the association between endometrial cancer and MAP remains contested because there is no statistically significant increase of endometrial cancer in MAP patients (12, 20, 55, 57-59). This association is based on three case reports. The first two describe a c.34G>T p.(Gly12Cys) transversion in the *KRAS* gene of MAP patients with endometrial cancer (59, 60). One of these patients developed endometrial cancer at 46, despite the average age of diagnosis being 61 in the general population. While the early onset of endometrial cancer has been associated with Lynch syndrome (59), this patient’s tumours were microsatellite stable and thus could not be the result of Lynch syndrome. The third case report described excess G:C>T:A transversions and the presence of the c.34G>T p.(Gly12Cys) variant in the *KRAS* gene in the endometrial cancer of a MAP patient (60). These observations could mean that there is a link between MAP and endometrial cancer, but investigations on a larger scale are required to fully validate this relationship (59).

Thyroid cancer has also been reported in MAP patients that have inherited at least one of the pathogenic founder variants (5, 12, 61-63). Although the association between thyroid cancer and FAP has been reported (12), it is difficult to describe thyroid cancer as one of the manifestations of MAP, especially in the absence of or the lack of information about Gly12Cys variants in the *KRAS* gene.

Pulmonary bronchioloalveolar carcinoma (BAC) is another type of cancer that has been reported in connection with MAP (64), although only in a single patient. In this case, molecular analysis identified G:C>T:A transversions in *KRAS* in both CRC samples and seven of nine pulmonary BAC samples. The presence of these transversions suggests that it could be part of the MAP phenotypic spectrum, but like the previously described cancers, a larger dataset is needed to definitively confirm any link between BAC and MAP.

Desmoids have been reported in three MAP patients (22, 52, 53). One of these patients had a mesenteric desmoid tumour, but the location of desmoids were not recorded for the others (53). The patient with the mesenteric desmoid tumour was homozygous for p.(Gly396Asp) and had an aggressive phenotype, characterised by over 100 tubular adenomas in the colon and 4 CRCs by the age of 30. Osteomas have only been described in four MAP patients to date (4, 52, 65). Because they are so rarely described in MAP, desmoids and osteomas might be distinguishing features between FAP and MAP (12).

Very few studies have reported Congenital Hypertrophy of the Retinal Pigment Epithelium (CHRPE) in MAP patients (4, 6, 12, 22, 38, 66), however it is unclear if these are the same type associated with FAP. In FAP patients, CHRPE is characterized by multiple bilateral, diffusely distributed, sharp bordered lesions. Sebaceous gland adenomas have also been reported multiple times in the literature (12, 39, 58, 61, 67, 68), which suggests that its association with colorectal cancer is not confined to patients with Lynch syndrome (12). Other benign extraintestinal manifestations that have been reported in MAP patients thus far include dental cysts, dermoid cysts, jawbone cysts, hepatic cysts, kidney cysts, lipoma, benign endometrial tumours and benign breast tumours (2, 4, 6, 12, 65).

## 4. Conclusion

While a phenotypic correlation with specific pathogenic variants was not observed, the founder variants carry an increased risk of colorectal cancer (6, 33, 34). When compared to p.(Gly396Asp), the variant p.(Tyr179Cys) is associated with increased CRC and duodenal adenoma risk, earlier onset, and more severe phenotype. The term *MUTYH*-associated polyposis implies that polyps are an obligatory phenotype, but numerous CRC cases in the absence of polyps suggests otherwise. There are also a multitude of extraintestinal manifestations associated with MAP or seen in MAP patients. Bladder, skin, ovarian and duodenal cancers, and adenomas have been found in multiple MAP patients, but the increased risk of developing endometrial, breast and thyroid cancer in these patients remains controversial. Desmoids and osteomas are rarely described in MAP, and thus might be features that distinguish MAP from FAP. There are several other benign extraintestinal manifestations reported in the literature, but their association with MAP has yet to be concretely established.

According to the findings of this narrative review, we would like to suggest the adoption of “*MUTYH*-associated tumour syndrome,” a term first described in Magrin et al. (2022) (11). This new definition of MAP encompasses the extracolonic phenotypic spectrum caused by *MUTYH* deficiency while excluding the presence of polyps as a necessary criterion for diagnosis. Because there is an overlap between the clinical and molecular characteristics of *MUTYH*-associated tumour syndrome and Lynch syndrome, underlying constitutional variants in mismatch repair genes must be ruled out before considering *MUTYH* deficiency as potential diagnosis. The somatic transversion that leads to the Gly12Cys variant in *KRAS* and *APC* is a well-documented consequence of *MUTYH* deficiency and could be used to firmly establish a causal relationship between MAP and the controversial phenotypes discussed here. This phenotypic review will facilitate the work of the InSiGHT-ClinGen Variant Curation Expert Panel as it outlines important phenotypic features of MAP for assessing variant pathogenicity, and ultimately contribute to improved patient care.

## Data Availability

All data produced in the present work are contained in the Supplementary section of the manuscript.

## List of abbreviations

ACMG: American College of Medical Genetics and Genomics
AMP: American Association of Molecular Pathology
APC: Adenomatous Polyposis Coli
BER: Base Excision Repair
CHRPE: Congenital Hypertrophy of the Retinal Pigment Epithelium
CRC: Colorectal Cancer
FAP: Familial adenomatous polyposis
HP: Hyperplastic Polyps
MAP: *MUTYH*-associated polyposis
MMR: Mismatch Repair
SIR: Standardized Incidence Ratio
SSL: Sessile Serrated Lesions

## Acknowledgements

This work was supported by NIH grant number U24CA258119 (PI Marc Greenblatt). John-Paul Plazzer was also supported by a subaward of this grant and by the Alan Watt and Chris Geyer Oncology Fellowship through The Royal Melbourne Hospital Foundation. Gabriel Capella is supported by the Spanish Ministry of Science and Innovation, which is part of Agencia Estatal de Investigación (AEI), through the Retos Investigación grant (PID2019-111254RB-I00/ DOI: 10.13039/501100011033). This study was also supported by CIBERONC (CB16/12/00234) and IMP/00009 financed by the Instituto de Salud Carlos III (ISCIII) and Fondo Europeo de Desarrollo Regional (FEDER).

## Data Availability

Data is provided in the supplementary section.

## Supplemental Section

**Table S1.**
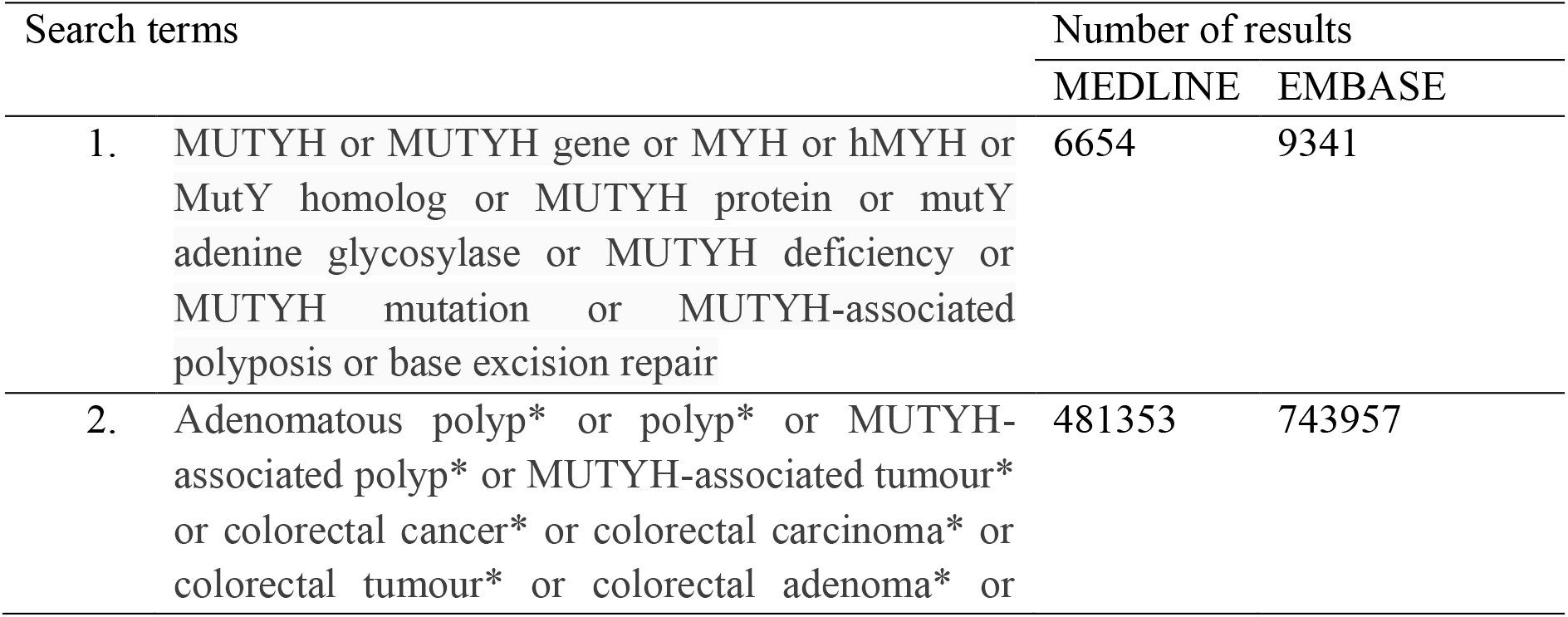
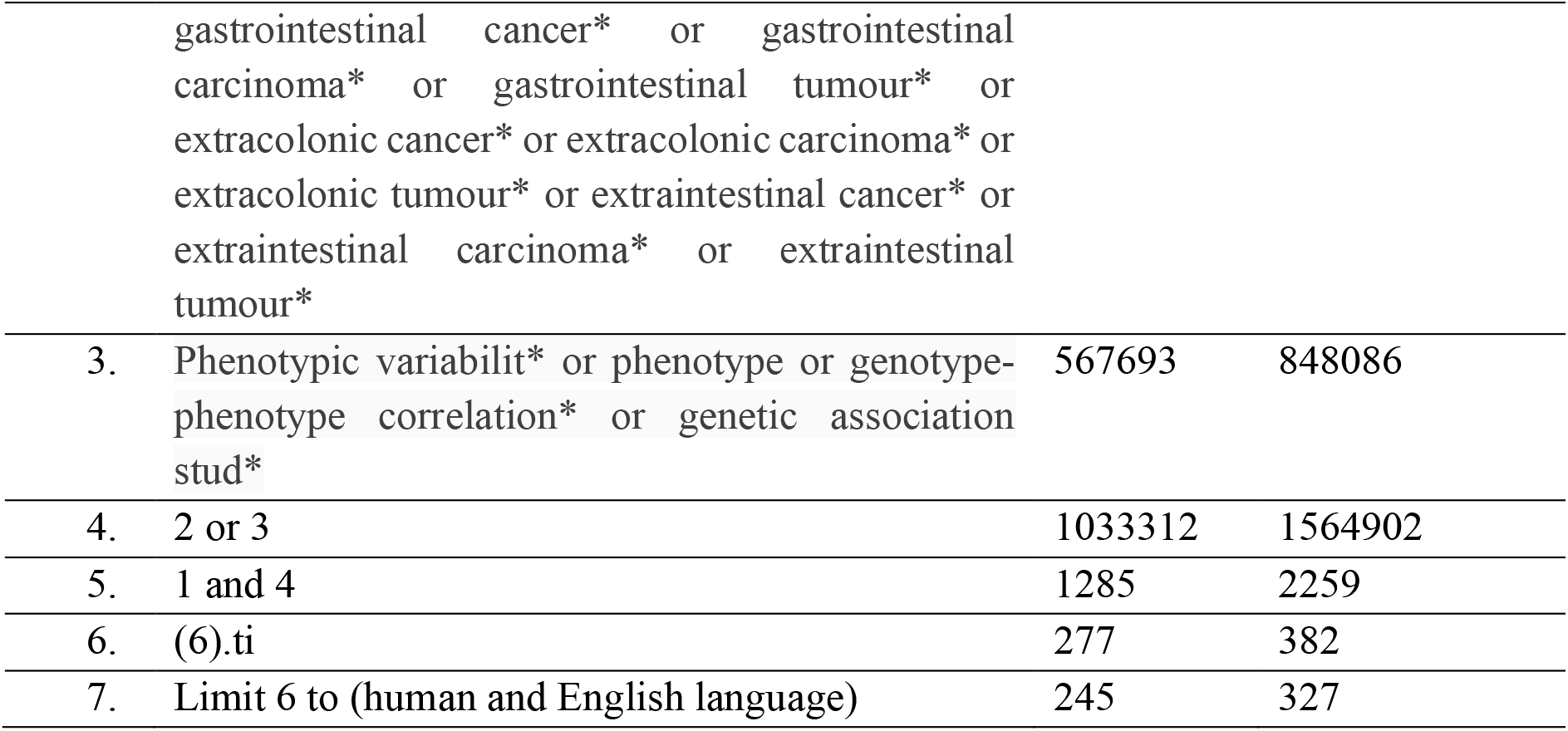
Research Strategy and Results (6/6/2022 – 15:40 AEST).

| Search terms | Number of results |  |
| --- | --- | --- |
|  | MEDLINE | EMBASE |
| 1. MUTYH or MUTYH gene or MYH or hMYH or MutY homolog or MUTYH protein or mutY adenine glycosylase or MUTYH deficiency or MUTYH mutation or MUTYH-associated polyposis or base excision repair | 6654 | 9341 |
| 2. Adenomatous polyp* or polyp* or MUTYH-associated polyp* or MUTYH-associated tumour* or colorectal cancer* or colorectal carcinoma* or colorectal tumour* or colorectal adenoma* or | 481353 | 743957 |

|  |  |  |  |
| --- | --- | --- | --- |
|  | gastrointestinal cancer* or gastrointestinal carcinoma* or gastrointestinal tumour* or extracolonic cancer* or extracolonic carcinoma* or extracolonic tumour* or extraintestinal cancer* or extraintestinal carcinoma* or extraintestinal tumour* |  |  |
| 3. | Phenotypic variabilit* or phenotype or genotype-phenotype correlation* or genetic association stud* | 567693 | 848086 |
| 4. | 2 or 3 | 1033312 | 1564902 |
| 5. | 1 and 4 | 1285 | 2259 |
| 6. | (6).ti | 277 | 382 |
| 7. | Limit 6 to (human and English language) | 245 | 327 |

**Table S2.**
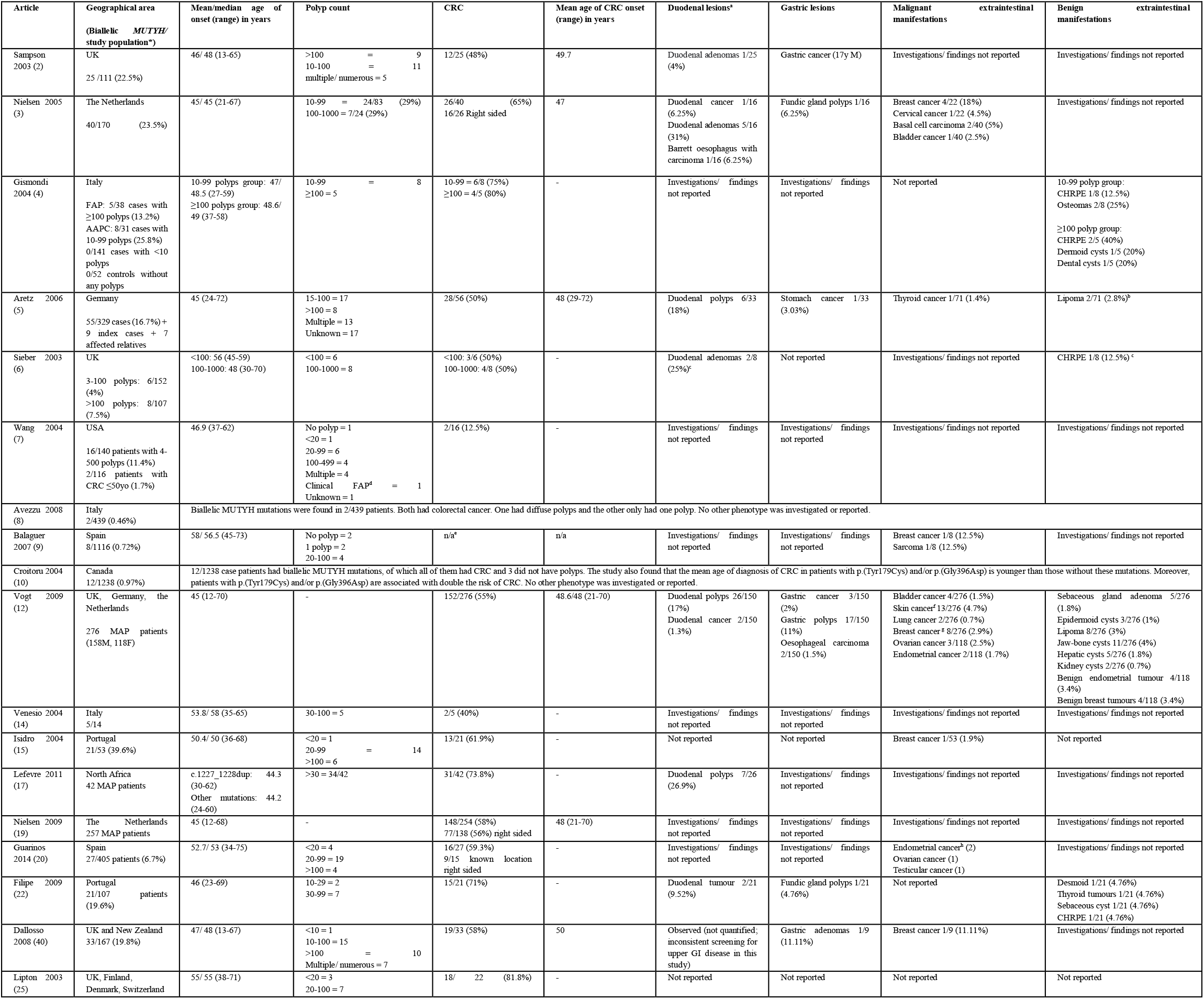
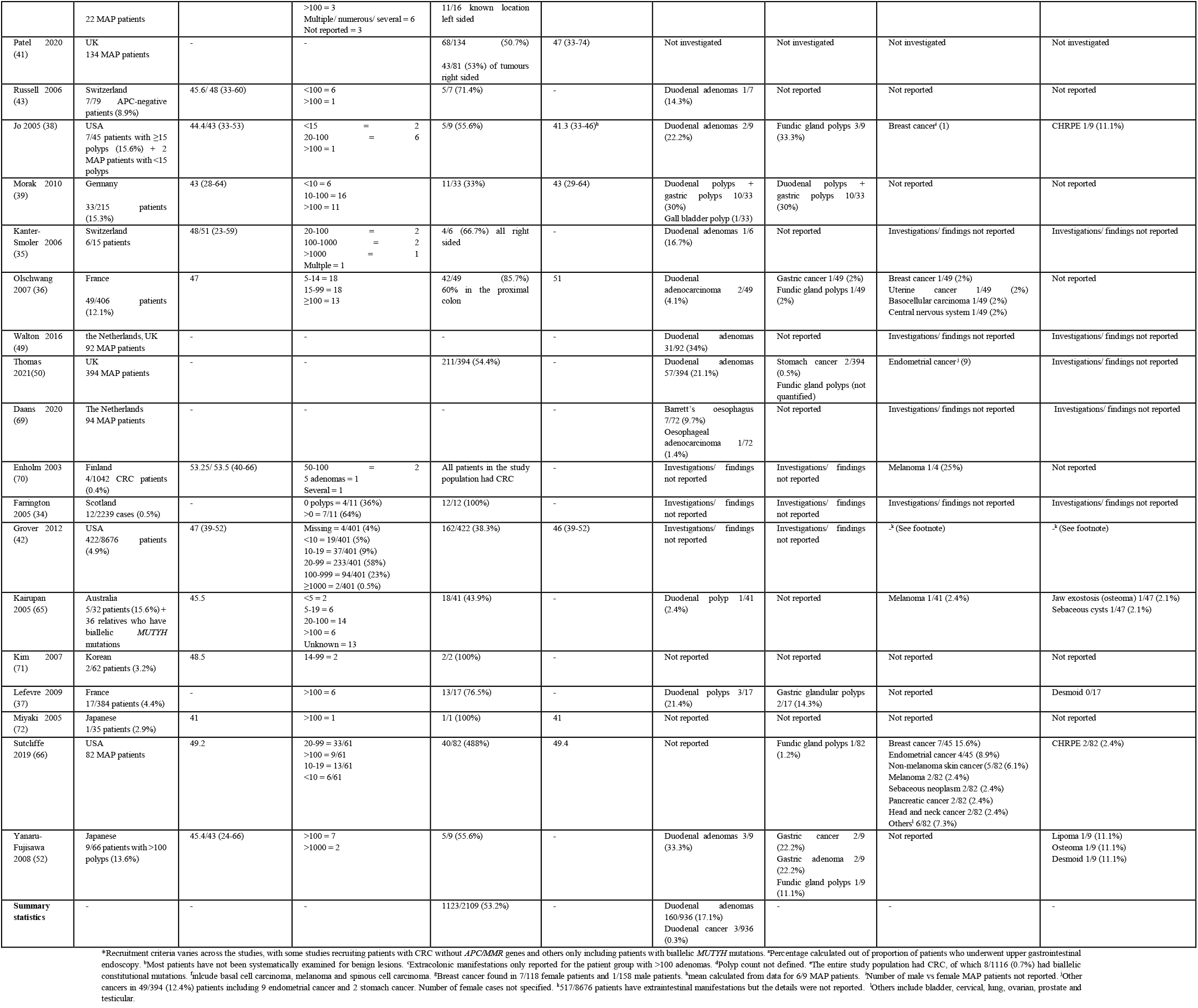
Clinical Features of Patients with Biallelic MUTYH Mutations.

In this table, “not reported” was used if a study investigated for a particular phenotype that was not found in the patients, “not investigated” was used if it was explicitly stated that the phenotype was not investigated, and “investigations/findings not reported” if there was no mention of investigation of the phenotype in the study.

